# Age-related changes in acoustic cue use for speech-in-speech perception

**DOI:** 10.64898/2026.06.17.26355866

**Authors:** Elizabeth Fish, Mishaela DiNino

## Abstract

Acoustic cues such as pitch and spatial location allow listeners to attend to a target speaker and ignore competing talkers, aiding speech recognition in background noise. Diminished ability to utilize acoustic cues for speech stream segregation may thus contribute to older adults’ challenges hearing in noise. Adults aged 18-74 completed a speech-in-speech identification task with three conditions containing 1) only pitch cues (fundamental frequency), 2) only spatial cues (interaural time differences; ITDs), or 3) both pitch and spatial cues for segregating a target talker from competing talkers. Hearing thresholds at standard and extended high frequencies (EHFs), auditory brainstem responses (ABRs), and digit span scores were acquired to examine whether sensory and cognitive factors that change with age influence acoustic cue use for speech-in-speech recognition. Significant differences were observed between cue condition scores indicating that use of the available cue(s) drove performance. Older adults (aged 40+) performed significantly more poorly on all cue conditions relative to younger adults. Hearing thresholds at standard frequencies significantly predicted scores on the Pitch condition. EHF hearing thresholds better predicted Spatial and Both Cue condition performance. ABR metrics were not a predictor in the expected direction but backward digit span scores significantly predicted scores on the more difficult conditions, demonstrating greater cognitive resource reliance in challenging listening scenarios regardless of acoustic content. Age-related changes in auditory function may therefore impair older adults’ speech-in-noise perception by reducing their ability to use acoustic cues for segregating target and competing speech.

## 1. Introduction

Perceiving speech in background noise involves attending to target speech while ignoring competing sounds. Various acoustic cues, such as differences in spatial location and in frequency between the target and maskers, enable listeners to successfully identify and focus on a target when other sounds are present (Woods et al., 2001; Shinn-Cunningham, 2008). Physiological sensory coding and higher-level perception, which inherently involves cognitive processes, both contribute to one’s ability to utilize acoustic cues for segregating targets from noise (Shinn-Cunningham, 2017). Yet, sensory acuity and cognitive function both decline with normal aging. Older individuals frequently report challenges hearing in background noise (CHABA, 1988; see Humes, 1996 for review), and it is thus possible that decreased ability to utilize the acoustic cues important for segregating speech from competing maskers contributes to these listening difficulties. Much research has examined age-related differences in speech-in-noise identification but less is known about how use of the acoustic cues themselves during speech-in-noise perception tasks changes with age.

For decades, research has demonstrated that the ability to hear speech in noise decreases as individuals age (e.g., Plomp & Mimpen, 1979). Older adults also find speech-in-noise perception more cognitively demanding compared to young adults (Desjardins & Doherty, 2013). Elevated pure tone thresholds are a normal consequence of aging; audiometric hearing loss provides one explanation for older adults’ challenges hearing in noise, as even young adults with hearing loss exhibit poorer speech-in-noise perception performance than their normal hearing peers (Souza & Turner, 1994; Dubno et al., 1984). Still, audiometric thresholds do not always predict a listener’s ability to understand speech in background noise, particularly in older adults (Kim et al., 2006). Previous studies have found that older adults have difficulty hearing speech in the presence of competing maskers even after controlling for hearing thresholds at 4 kHz and below (e.g., Frisina & Frisina, 1997). These findings indicate that other factors, such as audiometric thresholds at higher frequencies and/or age-related changes in neural processing, also play a role in older adults’ trouble hearing in the presence of competing sounds.

Precise auditory neural coding is important for perception of complex auditory signals but declines with normal aging. In young individuals, myelin actively regenerates, forming a sheath around axons to accelerate neuronal conduction times. However, myelin renewal decreases with age, slowing neural signal transmission (Rawji et al., 2023). An investigation of auditory neural changes from the brainstem to cortex found that older adults had reduced white matter integrity relative to young adults, consistent with age-related myelin degeneration (Fabrizio-Stover et al., 2025). Cochlear synaptopathy, the loss of synapses between inner hair cells and the auditory nerve, also occurs with age in humans (Wu et al., 2019). Fewer auditory nerve fibers transmitting auditory signals decrease the neuronal population response, impairing the precise phase-locking (and thus the auditory temporal processing) that is important for hearing in noise (Parthasarathy & Kujawa, 2018). Harris et al. (2021) also found age-related structural and functional changes in auditory nerve fibers, including reduced neural synchrony of phase-locking to auditory signals. In that study and in others (e.g., Anderson et al., 2011), lower neural synchrony significantly predicted poorer speech-in-noise recognition scores, demonstrating the importance of neural encoding for speech perception in background noise.

One metric of auditory neural function is the auditory brainstem response (ABR), an electrophysiological measure that assesses both neural transmission speed (assessed by wave latency) and the population response (assessed by wave amplitude) of neurons that form the auditory nerve and auditory brainstem nuclei (Jewett & Williston, 1971). ABRs are measured noninvasively through electrodes on the scalp, typically in response to broadband clicks to activate large numbers of hair cells and thus the auditory neurons that receive signals from those hair cells. ABRs can provide important information about the auditory pathway and are commonly used in clinical settings. Many investigations have also used ABRs to examine the effects of aging or hearing loss on neural transmission of sound. Age-related changes in ABR amplitudes and latencies are well-established, mirroring the neuronal changes that occur as individuals age. Wave latencies increase, reflecting slower transmission of auditory signals, and wave amplitudes decrease, reflecting fewer numbers of responding neurons, in older adults regardless of the degree of hearing loss (Konrad-Martin et al., 2012). Aging has been shown to affect wave V, the most robust ABR wave, even in normal hearing adults (Jerger and Hall, 1980).

Previous research suggests that auditory neural function begins to decrease even earlier than what is typically considered “older adulthood.” In a postmortem study, Kumar (2022) found that adults aged 31-50 demonstrated thinner myelin sheaths and smaller diameter of auditory nerve fibers compared to younger adults, with even further reduction in myelin and axon size in an older adult group. Consistent with these physiological findings, Jerger and Hall (1980) showed that average ABR wave V latencies were slightly increased in participants aged 30-39 relative to participants aged 20-29, with a much larger increase in latency between participants aged 30-39 and those aged 40-49. Zink et al. (2025) obtained envelope following responses (EFRs), a measure of subcortical neuron phase-locking to sound, in adults with normal hearing and found that the middle-aged adults (aged 40-55) had significantly reduced EFRs relative to young adults. In addition, age-related decreases in EFRs demonstrated in middle-aged human participants were similar to those in gerbils with confirmed age-related neural degeneration.

Auditory neuronal changes in middle-age are not limited to peripheral neuron and brainstem processing: Guo et al. (2025) measured cortical responses to phonemes in continuous speech and found “blurrier” neuronal representations of the phonemes in middle-aged (aged 40-54) than in younger adults, suggesting less accurate cortical tracking and coding of speech sounds in the middle-aged group. Therefore, age-related changes in auditory processing that are likely to affect speech-in-noise perception seem to begin in middle age. Trouble hearing in noise may also begin in middle age, as Zink et al. (2025) additionally found that middle-aged adults performed more poorly on a speech-in-noise recognition task compared to young adults, but only for the most difficult condition.

Evidence from previous investigations therefore indicates that age-related changes in sensory function can affect speech-in-noise understanding. This is a well-established relationship, but the mechanisms that link reduced sensory processing to challenges hearing speech in noise are not as clear. One potential explanation is that reduced sensory processing decreases the coding fidelity of the acoustic cues that contrast a target talker from competing sounds. For example, the decrease in outer hair cell function associated with presbycusis reduces frequency selectivity due to broadened auditory filters (Peters & Moore, 1992), interfering with perception of spectral acoustic cues. Auditory neuron phase-locking weakens with age, hindering precise coding of auditory timing (Anderson et al., 2011; Harris & Dubno, 2017; Roque et al., 2019). Changes that occur with age in perception of spectral and temporal acoustic cues may thus be a connection between weaker sensory processing and impaired speech-in-noise perception.

Fundamental frequency (F0) is one cue that can help listeners differentiate a target talker’s voice from other talkers or background noise. F0 is primarily determined by the rate of vocal fold vibration, which depends on length, mass, and tension of the vocal folds (Jiang et al., 2000) and therefore differs from person to person. Previous research has found that young, normal hearing adults perform better on speech recognition in the presence of a competing talker when the target and competing talker are of different sexes – and therefore have very different F0s (Brungart, 2001; Kidd et al., 2016). Other studies have demonstrated improvements in speech-in-speech identification with increased F0 differences between target and masker (Brokz & Noteboom, 1982; Assmann, 1999; Drullman & Bronkhorst, 2004). F0 is therefore a robust cue for perceiving speech in background noise. However, perception of spectral cues for differentiating speech sounds decreases with age (DiNino, 2024; Souza et al., 2011) and older adults may be less able than young adults to utilize F0 for segregating target speech from background speech.

Target talkers are typically in a different spatial location in the environment relative to distractors; consequently, spatial cues are important for accurate speech perception in noise. Spatial separation from target and masker also results in “spatial release from masking,” an improvement in performance relative to when both speech and background noise are in the same location (see Litovsky, 2012 for review). To achieve spatial release from masking, however, listeners must perceive and utilize the available spatial cues (Gallun et al., 2005; Gallun et al., 2013). Interaural time differences (ITDs) are the small differences in the time it takes for a sound to reach the left and right ears based on its location relative to the listener and are the primary cue for sound location in the horizontal plane (Wightman & Kistler, 1992). Prior investigations have shown that ITD perception (Strouse et al., 1998; Grose & Mamo, 2010) and spatial release from masking (see Koehnke & Besing, 2011 for review) both decline with age. Challenges in using ITDs to locate a target talker and differentiate it from a masker may hinder older adults’ speech-in-noise understanding.

Acoustic cue perception can be measured with simple detection or discrimination tasks, but those tasks do not provide information about how cues are used to segregate speech from competing speech. In contrast, speech-in-noise perception tasks which emphasize auditory selective attention require listeners to rely heavily on the acoustic characteristics that differentiate the target and masker sounds to distinguish target speech from maskers (Shinn-Cunningham, 2008). These tasks also tend to minimize linguistic contributions to task performance by using streams of digits or phonemes, as opposed to words or sentences, as the speech stimuli. Speech-in-speech perception tasks with these characteristics are more sensitive to basic sensory (e.g., Ruggles et al., 2011) and perceptual (Bharadwaj et al., 2015; Parthasarathy et al., 2020) processes than are more commonly used tests of sentence or word recognition in noise (see also DiNino et al., 2021).

This study therefore used streams of phonemes as “speech” to design a speech-in-speech recognition test that limited higher-order cognitive processing of syntax and word meaning while emphasizing reliance on acoustic cues to segregate a target speech stream from competing streams. Target and distractors were differentiated by F0 only, spatial location (using ITDs) only, or both F0 and ITDs to determine young and older adults’ use of acoustic cues for speech-in-speech perception, with the hypothesis that this ability would decrease with age. We hereafter refer to “cue use” as reflections of performance on each cue condition of the task rather than basic cue detection or discrimination. Hearing thresholds at standard and extended high frequencies and ABRs were collected to examine the extent to which sensory processing at the cochlear and subcortical level contributed to participants’ ability to use F0 and ITDs in the speech-in-speech recognition task. Participants also completed a digit span task so that the contribution of working memory capacity to speech-in-speech recognition scores could be determined.

## 2. Methods

### 2.1 Participants

Ninety-eight adults were recruited from the University at Buffalo and surrounding community. Some participated for course credit and others for pay. Seven individuals did not meet hearing threshold criteria (≤ 50 dB HL from 250-4000 Hz) and did not continue the study but were compensated for their time. Seven participants needed to be reinstructed for the speech-in-speech task multiple times and/or performed at or below chance (33.3%) on more than one condition and therefore their data was excluded. The final participant sample included 84 individuals aged 18 – 74 (mean age = 39.92, SD = 19.92 years). Participants gave written informed consent prior to completing any study tasks. All experimental procedures were approved by the University at Buffalo Institutional Review Board (STUDY00007816).

### 2.2 Audiometric Assessment

Otoscopy was first performed to visualize the tympanic membrane and ensure that participants did not have an ear infection or ear canal occlusion that would either disrupt perception or make earphone insertion unsafe. Pure tone audiometry was then conducted using Interacoustics Affinity Suite for standard frequencies (250-8000 Hz in octaves) and extended high frequencies (EHFs; 10k, 12.5k, 14k, 16k Hz) using the Hughson-Westlake down-10, up-5 procedure. Participants sat in a chair in a sound-attenuating booth and heard tones played through RadioEar DD450 circumaural headphones. They pressed the button on a clicker when they heard the tone. A threshold was determined when the participant responded that they heard the tone at least two out of three times that it was presented. Thresholds at each frequency were averaged across ears to obtain one bilateral threshold for each frequency, as the speech-in-speech identification task was presented through both ears simultaneously. For each participant, thresholds from 250-8000 Hz were averaged to yield a “standard frequency” threshold metric.

This frequency range was chosen to match the range tested in standard audiometric threshold evaluation and because the speech-in-speech perception task stimuli contained strong frequency energy up to ∼7900 Hz. Many older participants were unable to hear the EHF tones at 14kHz and 16kHz and therefore only thresholds from 10 kHz and 12 kHz were averaged to create an “EHF threshold” metric. These values were used in statistical models to examine the contribution of hearing thresholds at standard and extended high frequencies on speech-in-speech recognition scores.

Auditory brainstem responses (ABRs) were recorded with Intelligent Hearing Systems Smart EP to examine the contributions of subcortical neuron population responses to participants’ use of ITD and F0 cues for speech-in-speech perception. The clinical electrode configuration was used, with the non-inverting electrode placed below the hairline, the ground electrode between the eyebrows, and inverting electrodes placed on the left and right mastoids. Electrode placement sites were first cleaned with exfoliating gel and wiped off to optimize the electrodes’ contact with the skin. Electrode impedance levels were verified to be below 5 kΩ and were kept below 3kΩ whenever possible.

Participants were seated in a chair in a sound-attenuating booth that was then reclined during ABR collection to minimize muscle artifact. They wore ER-2 insert earphones and passively listened to a 90 dB nHL click with an alternating phase at a rate of 11.1/second to elicit the ABRs. Two runs containing 2048 sweeps were collected for each ear, totaling 4096 sweeps per ear. Following ABR collection, the examiner labelled waves I, III, and V and Smart EP automatically calculated the amplitude and latency for each wave. The speech-in-speech recognition task was presented binaurally and therefore ABR wave amplitudes and latencies were averaged across ears for each participant to create a bilateral ABR metric. One participant’s ABR data were excluded from analysis due to persistent artifact that could not be resolved with troubleshooting. This participant’s data for all other study metrics were included in statistical models.

ABR waves I and V were of particular interest in this study, as the neural generators for these two waves have been most commonly tied to speech understanding. Normalized ABR metrics based on both waves were used in the statistical models to limit 1) across-subject variability in neural integrity assessment and 2) multicollinearity, as raw ABR waves and amplitudes are often strongly correlated with hearing thresholds. The wave I/V amplitude was calculated for each participant and used as a metric of subcortical neural response strength. The inter-wave latency between wave V and wave I was calculated for each participant to create a metric of subcortical neural response speed. ABR wave I/V amplitude and wave V-I interpeak latency were included as independent variables in statistical analyses.

### 2.3 Study Tasks

#### 2.3.1 Speech-in-Speech Perception Task

Participants listened to “sentences,” hereafter referred to as streams, comprised of the syllables “ba,” “da,” and “ga” naturally spoken by a female talker. Streams of three phonemes, as opposed to stimuli with greater linguistic content such as words or sentences, were chosen as the speech stimuli to maximize the task’s sensitivity to basic sensory processing. Three streams each consisting of three random syllables were presented on each trial. Twenty combinations of “ba,” da,” and “ga” stimuli were predefined so that a stream could consist of two of the same syllables but never three of the same. Participants were instructed to focus on a target stream and ignore the other two streams.

Stimuli were recorded at a rate of 44.1 kHz. They were then bandpass filtered from 50 Hz to 10 kHz Hz using a pass Hann band with 100 Hz smoothing in Praat (Boersma, 2001) to remove recording noise. Visualization of the stimuli spectrograms indicated that frequency energy up to ∼7900 Hz remained in the stimuli. The stimuli were equated in duration so that participants could not use syllable duration as an identification cue for the phonemes in the target stream. Steps were also taken to minimize temporal overlap of the stop consonants in the three streams to allow for audibility. The phonemes were elongated from ∼350 ms each to 650 ms each using the “stretch” method in the “change duration” function in the Praat Vocal Toolkit (Corretge, 2012-2026) to allow for more stimulus timing options in which the stop consonants between streams did not overlap. The target stream always began first. The first distractor stream began 250 ms after onset of the target, followed 200 ms later by onset of the second distractor stream. The interstimulus intervals (ISIs) also differed between each phoneme in each stream, ranging from 250-400 ms. Figure 1 illustrates the stimuli timing on each trial. These timings were the same on each trial and were carefully chosen to reduce overlap between stimuli.

**Figure 1.**
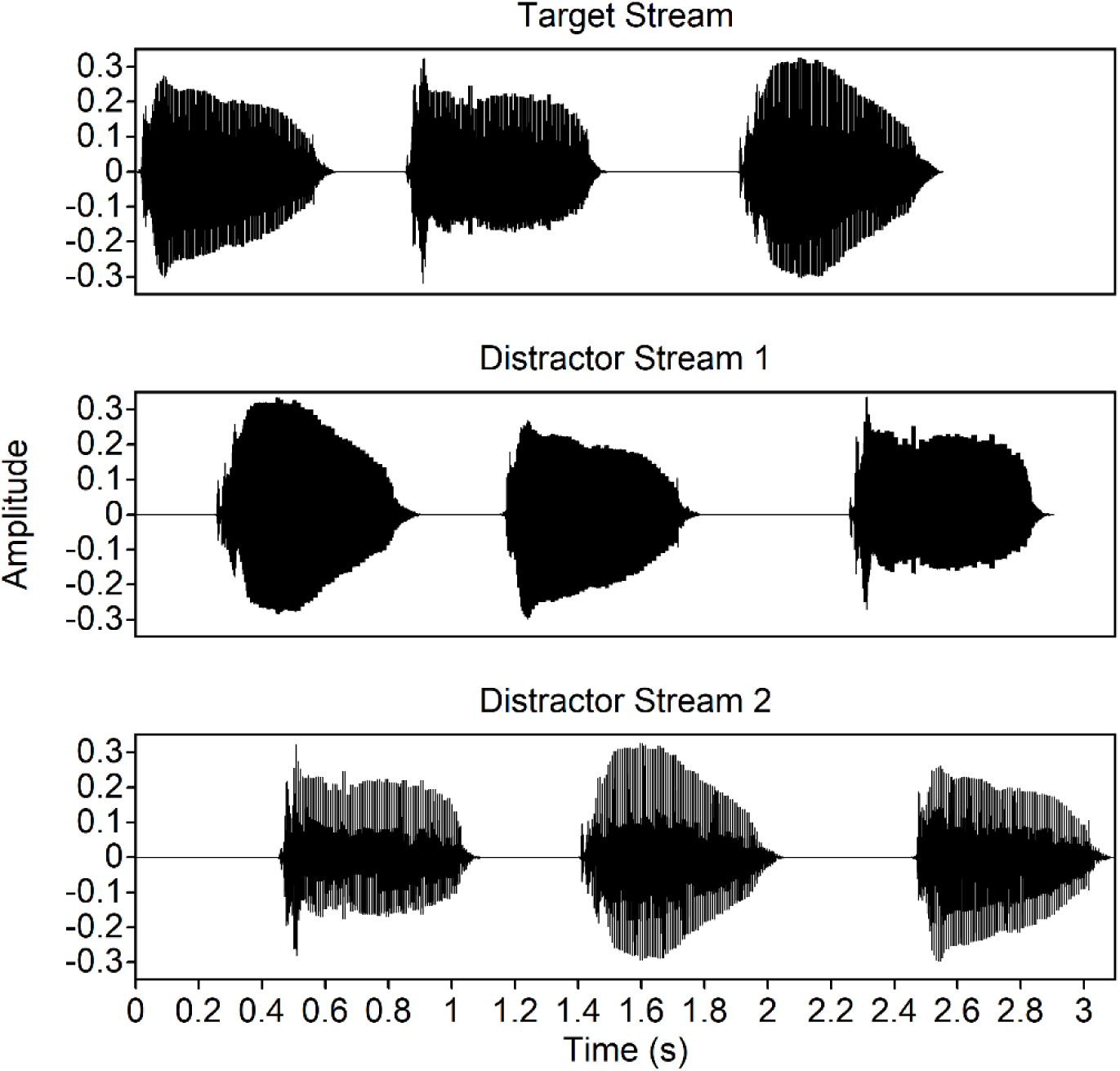
Timing of the three streams on an example trial in the speech-in-speech recognition task. Waveform amplitude is on the y-axis and time in seconds is on the x-axis. Onset timings and intervals between syllables within a stream were varied to minimize overlap so that the stop consonant at the beginning of each syllable was audible.

Variations in the ISI in each stream also facilitated streaming and better simulated the cadence of natural speech.

Characteristics of the target stream depended on the task condition. In the “Pitch” condition, the three streams were presented in the same spatial location (0°) but each differed in their F0. The “change semitones” function in the Praat Vocal Toolkit (Corretge, 2012-2026) was used to shift the F0 of the original stimuli (∼224 Hz) upward and downward by six semitones to create sets of “ba,” “da,” and “ga” syllables with a lower and a higher F0. The sound “eeeee,” spoken by the target talker (always the talker with an F0 of 224 Hz), was played at the beginning of each trial. Participants were instructed to listen to the “eeeee” presentation and then attend to the stream with the same F0 as the talker who said “eeeee.” In the “Spatial” condition, the target was presented at center (0°) and the distractor streams were spatially separated with a +/-300 µs ITD cue, simulating +/- 45° separation from center. This ITD was chosen to emphasize age effects, based on results from a previous investigation of ITD discrimination thresholds with age (Grose & Mamo, 2010). All streams were spoken by the same talker and therefore F0 cues were unavailable; ITDs were the only cue that a participant could rely on to identify the target stream. The “eeeee” cue was presented at the beginning of each trial in the Spatial condition to standardize task parameters, but as all talkers were the same in the Spatial condition, the F0 cue was not useful. Participants were instead instructed on each trial to attend to the stream presented at center.

In a third condition, termed “Both Cues,” streams were spatially separated with a +/- 600 µs ITD cue and also differed in their F0s to the same extent as in the Pitch condition. Therefore, participants could rely on either the ITD or F0 cue (again the talker who said “eeeee” at the beginning of the trial) to identify the target stream in the Both Cues condition. A larger ITD was used to increase the utility of the spatial cue, as pitch differences between streams in this study were relatively large (e.g., Darwin et al., 2003). Greater spatial separation between target and distractors in the Both Cues condition provided a better balance of usefulness between spatial and pitch cues.

Participants were seated in a sound-attenuating booth and wore ER-2 insert earphones to complete the speech-in-speech identification task. Audio was played at 70 dB SPL through an RME Fireface UFX III external sound card. Custom code written in MATLAB (The Mathworks, Inc.) using psychtoolbox functions (Kleiner et al., 2007) was used to present the task and record participant responses. The task was divided into six blocks (two for each condition) with each block containing 20 trials for a total of 120 trials. The order of blocks was completely randomized for each participant. On each trial, participants listened to the streams and used a computer mouse to click boxes labelled “Ba,” Da,” and “Ga” to indicate the syllables they heard, as well as the order that they heard them, within the target stream. Participants were told that they could take breaks in between blocks.

#### 2.3.2 Digit Span Task

Determining the role of sensory function in acoustic cue use for speech-in-speech perception was the primary interest in this study, but not all listening difficulties are related to auditory physiology. Participants performed a test of short-term working memory to determine the extent to which this cognitive ability influenced performance on the speech-in-speech perception task. A forward and backward digit span task was built using Gorilla Experiment Builder (www.gorilla.sc; Anwyl-Irvine et al., 2020). Digit sequences were presented visually on the computer screen. In the “forward” condition, participants were instructed to memorize the sequence and type the numbers back in the order they memorized into a response box at the end of the trial. In the “backward” condition, participants typed the digits in reverse order of their presentation. Each condition consisted of three practice trials in which sequences of three digits were presented. Following practice, the number sequences presented on each trial increased in length from three digits to four, five, six, and then seven. Participants completed three trials for each digit sequence length. Trials were scored as accurate if all presented digits on a trial were correctly identified. The percent of correct trials for the forward and backward conditions were calculated separately for each participant. Both variables were included in statistical analyses, as they reflect distinct cognitive processes: forward digit span measures memory capacity, whereas backward digit span requires both storage and manipulation of the digits, thereby also assessing executive function (e.g., Hale et al., 2002).

### 2.4 Statistical Analysis

All analyses were conducted in R software (R Core Team, 2025). Outlier analyses were first conducted with a Grubbs test using grubbs.test function in R. One datapoint in the ABR wave I/V ratio was identified as an extreme outlier and was excluded from statistical analyses and data visualizations. This participant’s data from other variables was maintained in the analyses. No other outliers were identified.

A mixed-model regression was conducted using the lmerTest package (Kuznetsova et al., 2017) to examine the effect of cue condition on participants’ scores on the speech-in-speech perception task. The regression model included task score as the dependent variable, cue condition as the independent variable, and random intercepts of overall score per participant to account for the same participants completing three task conditions. The Both Cue condition was set as the reference condition for analysis.

To examine whether age influenced scores on each cue condition of the speech-in-speech recognition task, participants were assigned to age groups and average scores on each cue condition were statistically compared between groups. Participants aged 18-39 (n = 50) were categorized as “younger” and those aged 40-74 (n = 34) were categorized as “older.” Age values for each group were based on previous research of auditory neural function in middle-aged adults (e.g., Jerger & Hall, 1981; Zink et al., 2025). Shapiro-Wilk tests using the shapiro.test function in R were first performed on speech-in-speech recognition test scores from each of the three conditions to determine whether the data were normally distributed, a requirement for t-test analyses. The results of all three tests indicated non-normality of the data (*p* < .05) and therefore Wilcoxon rank sum tests using the wilcox.test function in R were conducted to compare the cue condition score means between age groups.

Stepwise linear regressions were performed using the step() function in R to determine the variables that best predicted performance on each of the three conditions of the speech-in-speech perception task. Each model initially contained the independent variables of average standard frequency hearing threshold, average EHF hearing threshold, ABR I/V amplitude ratio, ABR wave V-I latency, forward digit span task score, and backward digit span task score.

Bidirectional stepwise regression analyses were then conducted to ascertain the best fitting model (the one that explains the greatest amount of variance in the dependent variable with the fewest number of dependent variables) for scores on the Pitch, Spatial, and Both Cue conditions. This analysis combined forward selection and backward elimination of variables to find the set of independent variables resulting in the lowest Akaike Information Criterion (AIC) value, an estimate of expected prediction error, for scores on each speech-in-speech task cue condition.

It was expected that many of the predictor variables in these models would cluster with age. The goal of the stepwise regression analyses was to examine the influence of factors that are known to change with age on use of acoustic cues for speech-in-speech recognition. Therefore, age was not included as a linear predictor in these models, as the intention was to determine the variance in performance explained by peripheral and cognitive factors and not age itself. This approach attempted to unpack the factors embedded in “aging” that could impact functional use of particular acoustic cues. However, correlation analyses using the cor.test function in R were run between age and the predictor variables to determine which effects yielded by the regression models were likely due to aging processes. Correction for multiple comparisons was not applied, as these analyses were conducted to determine each predictor variable’s unique relationship to age.

Transforming the speech-in-speech perception task scores to rationalized arcsine units (RAU) was explored to normalize error variance and mitigate any ceiling effects on task performance (Studebaker, 1985). The statistical model results using RAU values were very similar to those conducted with raw data and therefore the results of analyses using raw data are reported below.

## 3. Results

Mixed-model regression to examine the effect of cue condition on speech-in-speech identification scores revealed a significant effect of cue condition on task performance. Relative to the Both Cues condition, scores on the Pitch condition were significantly higher (*β* = 3.47, *p* = 0.027) and scores on the Spatial condition were significantly lower (*β* = -8.373, *p* < 0.001).

Distinct average scores between cue conditions suggests that participant’s use of the cue(s) available for speech stream segregation contributed to performance on the task. Figure 2 shows score distributions for each cue condition.

**Figure 2.**
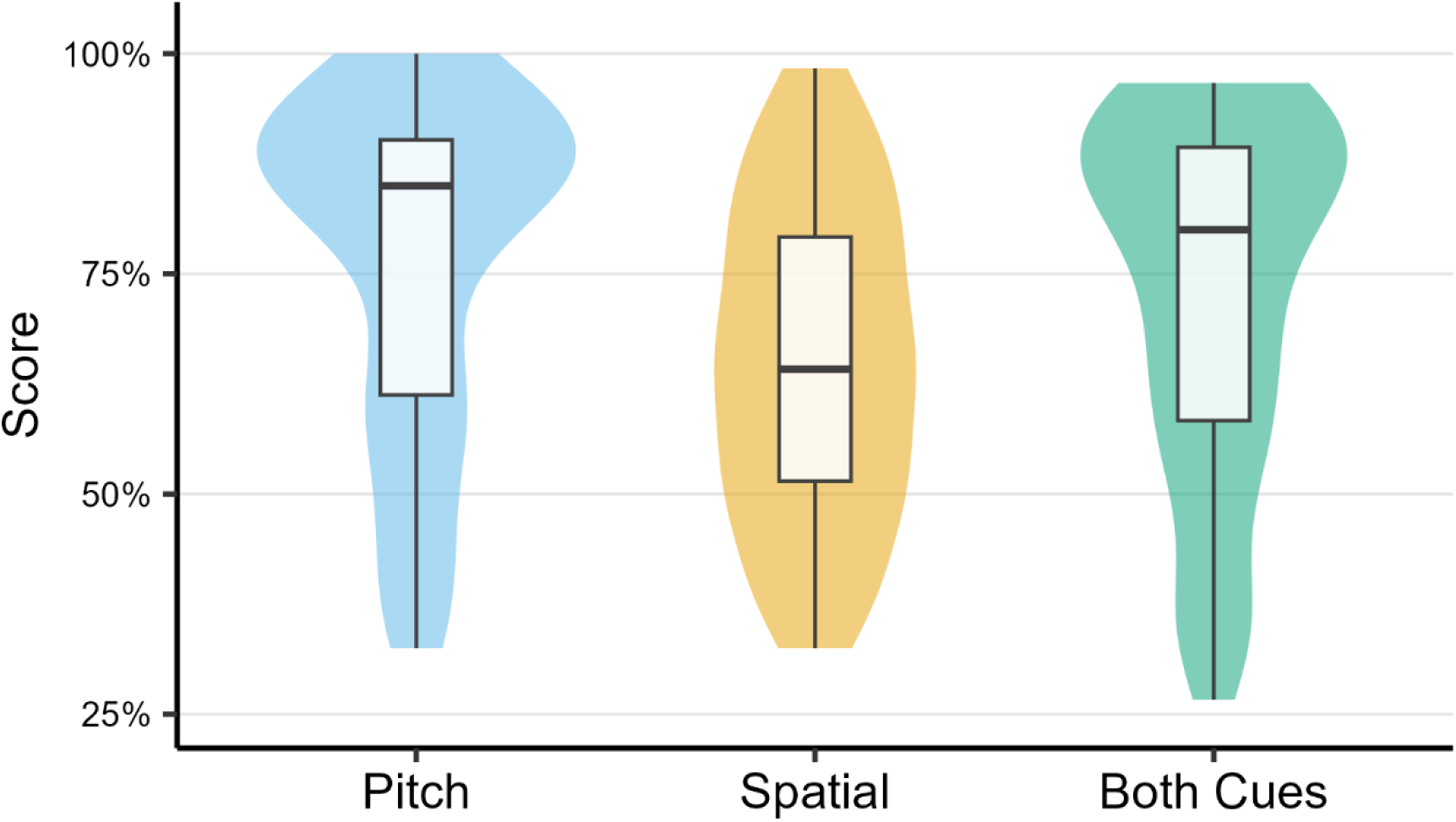
Distribution of scores, in percent correct, on the speech-in-speech perception task (y-axis) for each of the three cue conditions (x-axis). The shaded areas indicate score distribution density with wider sections representing higher density and narrower sections representing lower density. Boxplots are overlaid on score distributions from each cue condition. The top and bottom of each box represent the 75th and 25th quartiles and the line in the middle of each box represents the median. Whiskers extend to 1.5 times the interquartile range.

Age group analyses revealed significant differences on speech-in-speech recognition task performance between younger (aged 18-39) and older (aged 40+) participants. The older age group received significantly lower scores on the Pitch (*W* = 1267, *p* < 0.001), Spatial (*W* = 1289, *p* < 0.001), and Both Cues (*W* = 1360, *p* < 0.001) conditions. Figure 3 shows score distributions for each cue condition separated by age group.

**Figure 3.**
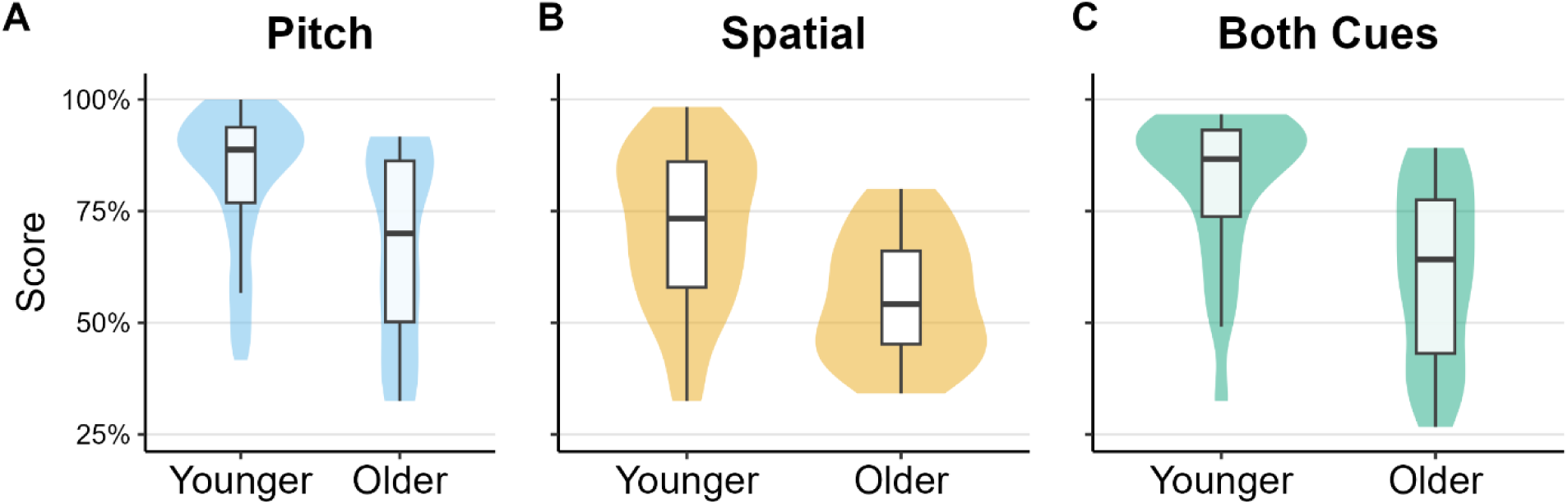
Score distributions for each cue condition (y-axes) separated by age group (x-axes). A) Pitch cue condition. B) Spatial cue condition. C) Both Cues condition. In all panels, the shaded areas indicate score distribution density with wider sections representing higher density and narrower sections representing lower density. Boxplots are overlaid on score distributions from each cue condition. The top and bottom of each box represent the 75th and 25th quartiles and the line in the middle of each box represents the median. Whiskers extend to 1.5 times the interquartile range.

As significant differences in speech-in-speech identification task performance were found between age groups, separate mixed-model regressions examining the effect of cue condition on task scores were performed with data from each group. The results of the analyses for the older adult group were consistent with results found in all data combined: scores on the Pitch condition were significantly higher (*β* = 5.809, *p* = 0.022) than scores on the Both Cues condition, and scores on the Spatial condition were significantly lower (*β* = -5.980, *p* = 0.019) than scores on the Both Cues condition. However, for younger adults, only scores on the Spatial condition were significantly different than those from the Both Cues condition (*β* = -10.00, *p* < 0.001). Pitch condition performance was statistically similar to performance when both cues were available (*β* = 1.883, *p* = 0.351).

Each participant’s score on the Spatial condition was subtracted from their score on the Pitch condition to generate a “performance difference” score for each participant, reflecting the degree to which individual listeners’ performance differed between conditions. A one-sample t-test was conducted on this metric to determine whether a statistically meaningful difference in performance existed between conditions, which would indicate that distinct processes, such as ability to use a particular cue, contributed to scores on each condition. Indeed, the t-test revealed that score differences between the Pitch and Spatial conditions were significantly different from zero [*t*(83) = 7.032, *p* < 0.001]. Figure 4 shows the change in performance between cue conditions among participants in both age groups.

**Figure 4.**
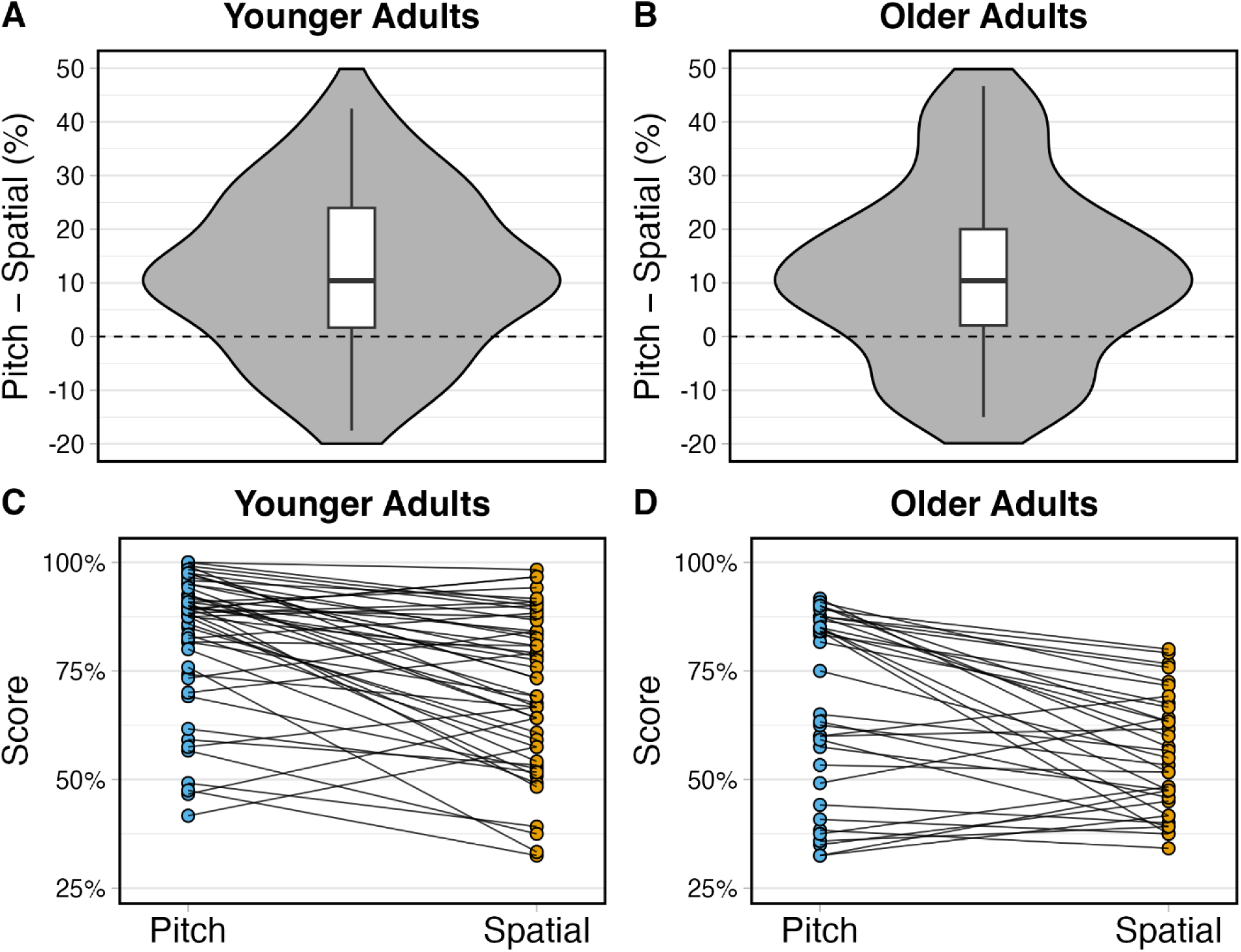
Differences in performance between the Pitch and Spatial cue conditions on the speech-in-speech recognition task. Top panels: Violin plots illustrating the distribution of the differences in scores between Pitch and Spatial cue conditions in percent correct (y-axes) for (A) younger and (B) older adults. In both panels, the shaded areas indicate score distribution density with wider sections representing higher density and narrower sections representing lower density. Boxplots are overlaid on score distributions from each cue condition. The top and bottom of each box represent the 75th and 25th quartiles and the line in the middle of each box represents the median. Whiskers extend to 1.5 times the interquartile range. Bottom panels: Line graphs depicting the differences in individual participant performance (y-axes) between the Pitch and Spatial cue conditions (x-axes) of the speech-in-speech recognition task for (C) younger and (D) older adults. Blue circles represent individual participants’ scores on the Pitch condition.

Yellow circles represent individual participants’ scores on the Spatial condition. The black line connects the Pitch and Spatial cue condition scores for each participant. Lines that slope download indicate that the participant achieved a higher score on the Pitch compared to the Spatial condition. Lines that slope upward indicate that the participant achieved a higher score on the Spatial compared to the Pitch condition.

Stepwise regression analysis was used to determine the participant factors that best predicted scores on each cue condition (and therefore use of each cue during speech stream segregation) in the speech-in-speech perception task. The variables that comprised the best-fitting models and their statistical effects are shown in Table 1. Average hearing thresholds at standard frequencies significantly predicted scores on the Pitch condition (see Figure 5A). The inclusion of forward and backward digit span scores improved model fit for the Pitch condition but these effects were not significant (*p* > 0.05; see Table 1). EHF hearing thresholds (see Figure 5B) and backward digit span task scores (see Figure 6B) were significant predictors of performance on the Spatial and the Both Cues conditions. ABR wave I/V amplitude was also significant predictor of Spatial condition performance but the effect was in the opposite direction than anticipated: individuals with larger ABR wave I/V ratios, reflecting greater auditory nerve responses, performed more poorly on the task (see Figure 5C). ABR wave V-I interpeak latency was not a significant predictor of scores on any cue condition (see Figure 5D).

**Figure 5.**
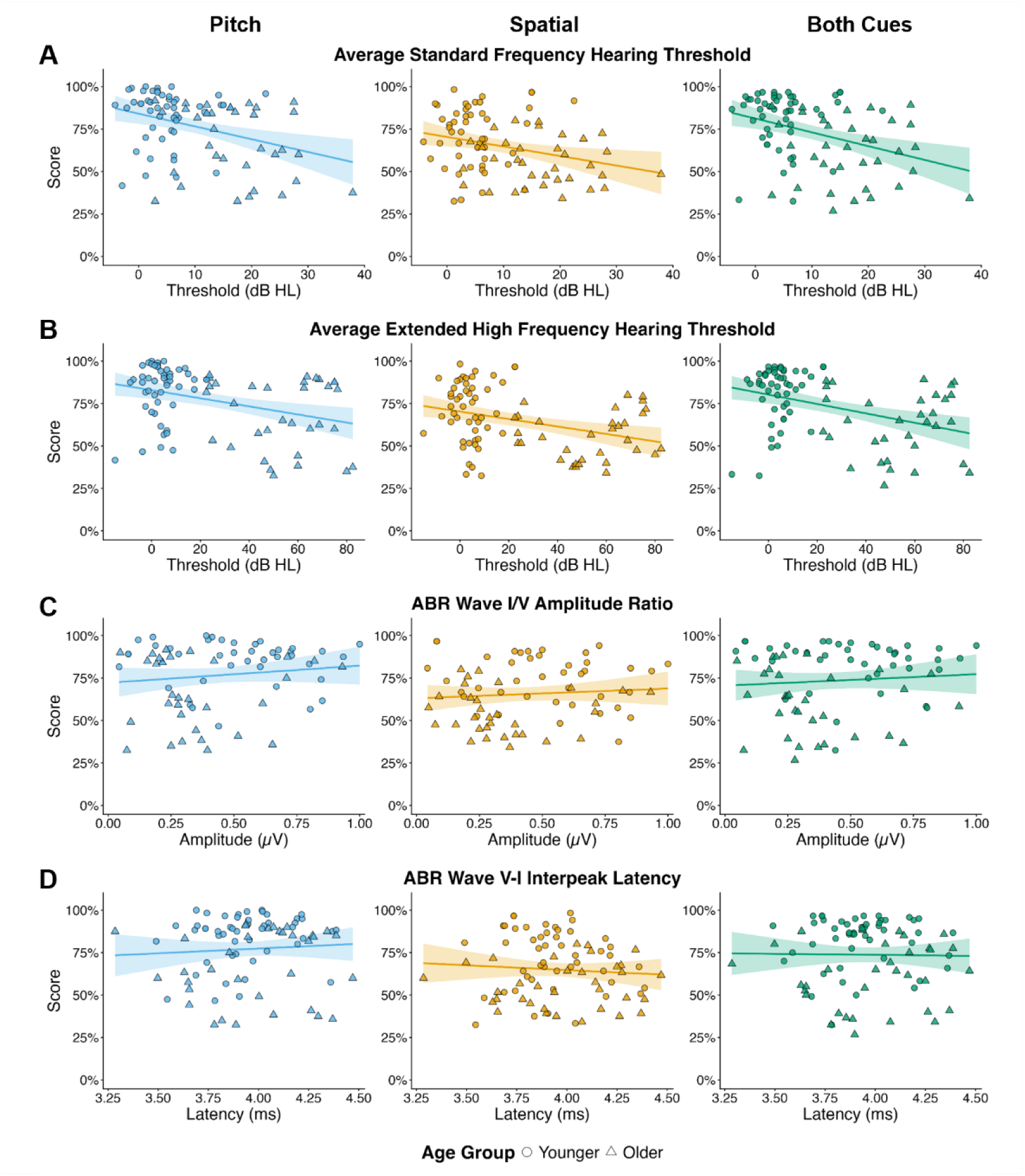
Relationships between sensory predictor variables (x-axes) and participant scores in percent correct (y-axes) from each cue condition of the speech-in-speech identification task. Predictor variables include A) hearing thresholds at standard frequencies (250-8000 Hz), B) hearing thresholds at extended high frequencies (10 and 12.5 kHz), C) auditory brainstem response wave I/V amplitude ratio, and D) auditory brainstem response wave V-I interpeak latency. Speech-in-speech identification task cue conditions include Pitch (left panels), Spatial (middle panels), and Both Cues (right panels). The line on each plot indicates the line of best fit, with the shaded area around the line representing the 95% confidence interval. Circles represent data from individual younger adults and triangles represent data from individual older adults.

**Figure 6.**
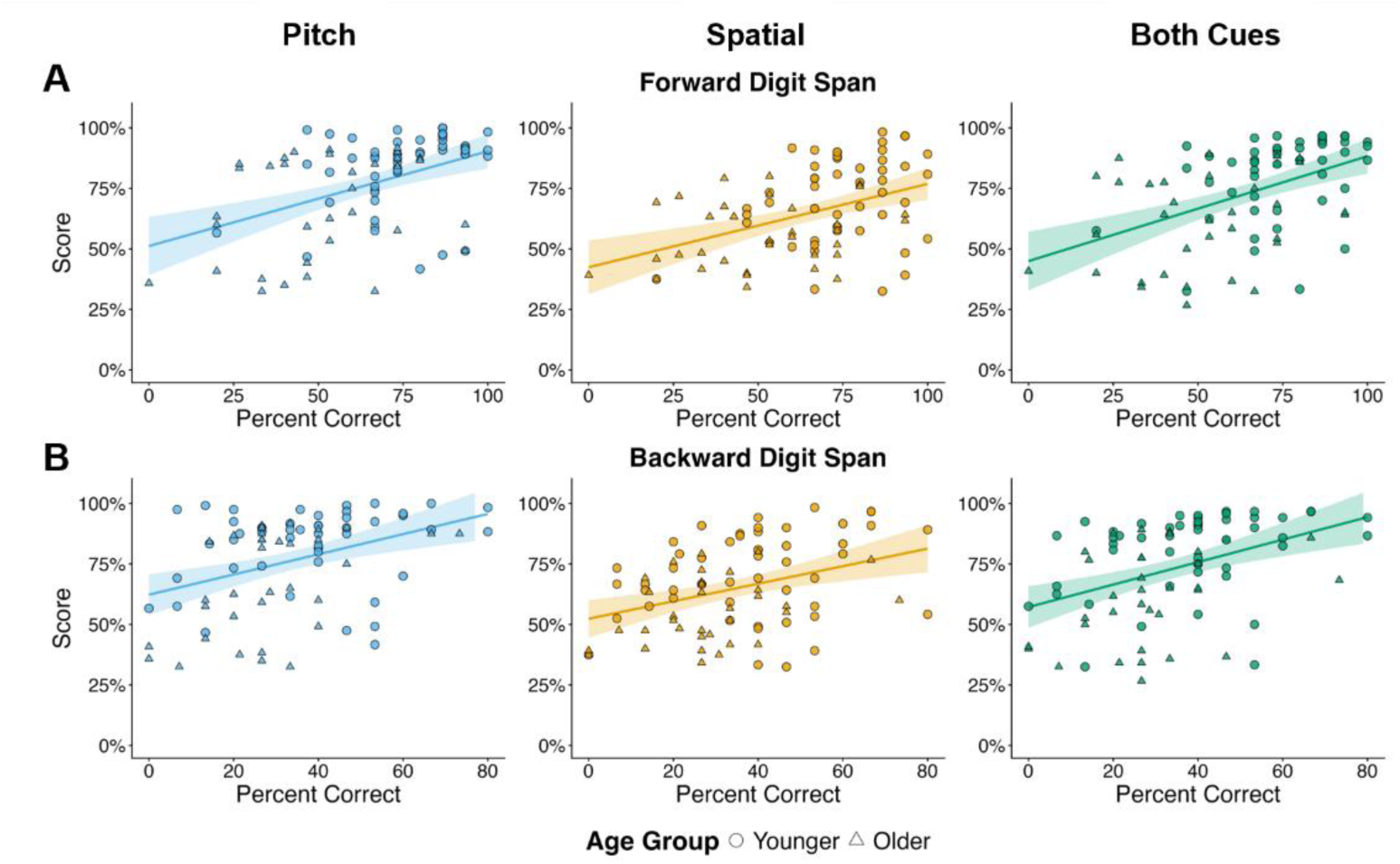
Relationships between digit span task scores in percent correct (x-axes) and participant scores in percent correct from each cue condition of the speech-in-speech identification task (y-axes). Predictor variables include A) forward digit span task scores, reflecting memory span, and B) backward digit span task scores, reflecting mental flexibility and executive function. Speech-in-speech identification task cue conditions include Pitch (left panels), Spatial (middle panels), and Both Cues (right panels). The line on each plot indicates the line of best fit, with the shaded area around the line representing the 95% confidence interval. Circles represent data from individual younger adults and triangles represent data from individual older adults.

**Table 1.** Statistics from the best-fitting stepwise regression models. The combination of variables shown for each cue condition of the speech-in-speech recognition task were revealed as the best predictors of scores on that condition. Note that some variables improved predictive model fit but were not significant predictors.

| Cue Condition | Model Fit |  |  | Best Fitting Model Statistics |  |  |  |  |
| --- | --- | --- | --- | --- | --- | --- | --- | --- |
| | *F* | df | *p* | Factor | $\beta$ | SE | *t* | *p* |
| Pitch | 10.87 | 3, 77 | < 0.001 | Intercept | 61.378 | 7.404 | 8.290 | <0.001* |
|  |  |  |  | Standard hearing threshold | -0.506 | 0.232 | -2.181 | 0.032* |
|  |  |  |  | Forward digit span | 0.204 | 0.115 | 1.779 | 0.079 |
|  |  |  |  | Backward digit span | 0.231 | 0.126 | 1.820 | 0.073 |
| Spatial | 10.2 | 3, 77 | < 0.001 | Intercept | 66.097 | 5.133 | 12.877 | < 0.001* |
|  |  |  |  | EHF hearing threshold | -0.248 | 0.067 | -3.722 | < 0.001* |
|  |  |  |  | ABR wave I/V ratio | -13.430 | 5.68 | -2.364 | 0.021*† |
|  |  |  |  | Backward digit span | 0.359 | 0.098 | 3.646 | < 0.001* |
| Both Cues | 15.49 | 3, 77 | < 0.001 | Intercept | 72.586 | 5.156 | 14.077 | < 0.001* |
|  |  |  |  | EHF hearing threshold | -0.311 | 0.067 | -4.641 | <0.001* |
|  |  |  |  | ABR wave I/V ratio | -10.774 | 5.707 | -1.888 | 0.063† |
|  |  |  |  | Backward digit span | 0.438 | 0.099 | 4.433 | <0.001* |

\* $p < 0.05$
† Relationship is in the opposite direction than expected.
SE = standard error
df = degrees of freedom
ABR = auditory brainstem response
EHF = extended high frequency

Correlation analyses revealed that all predictor variables except ABR wave V-I latency changed significantly with age. Table 2 shows the results of the correlations. Figure 7 shows the relationship between each predictor variable and participant age. These findings suggest that age drives changes in sensory and cognitive factors that then influence processes necessary for speech-in-speech recognition.

**Table 2.** Statistical results of the correlation analyses conducted between predictor variables and participant age. Positive *R* values indicate that the variable increased with older age. Negative *R* values indicate that the variable decreased with older age.

| Predictor Variable | $R$ | $p$ |
| --- | --- | --- |
| Standard hearing thresholds | 0.723 | < 0.001* |
| EHF hearing thresholds | 0.901 | < 0.001* |
| ABR wave I/V ratio | -0.333 | 0.002* |
| ABR wave V-I latency | 0.124 | 0.262 |
| Forward digit span score | -0.445 | < 0.001* |
| Backward digit span score | -0.236 | 0.031* |

\* $p < 0.05$
ABR = auditory brainstem response
EHF = extended high frequency

**Figure 7.**
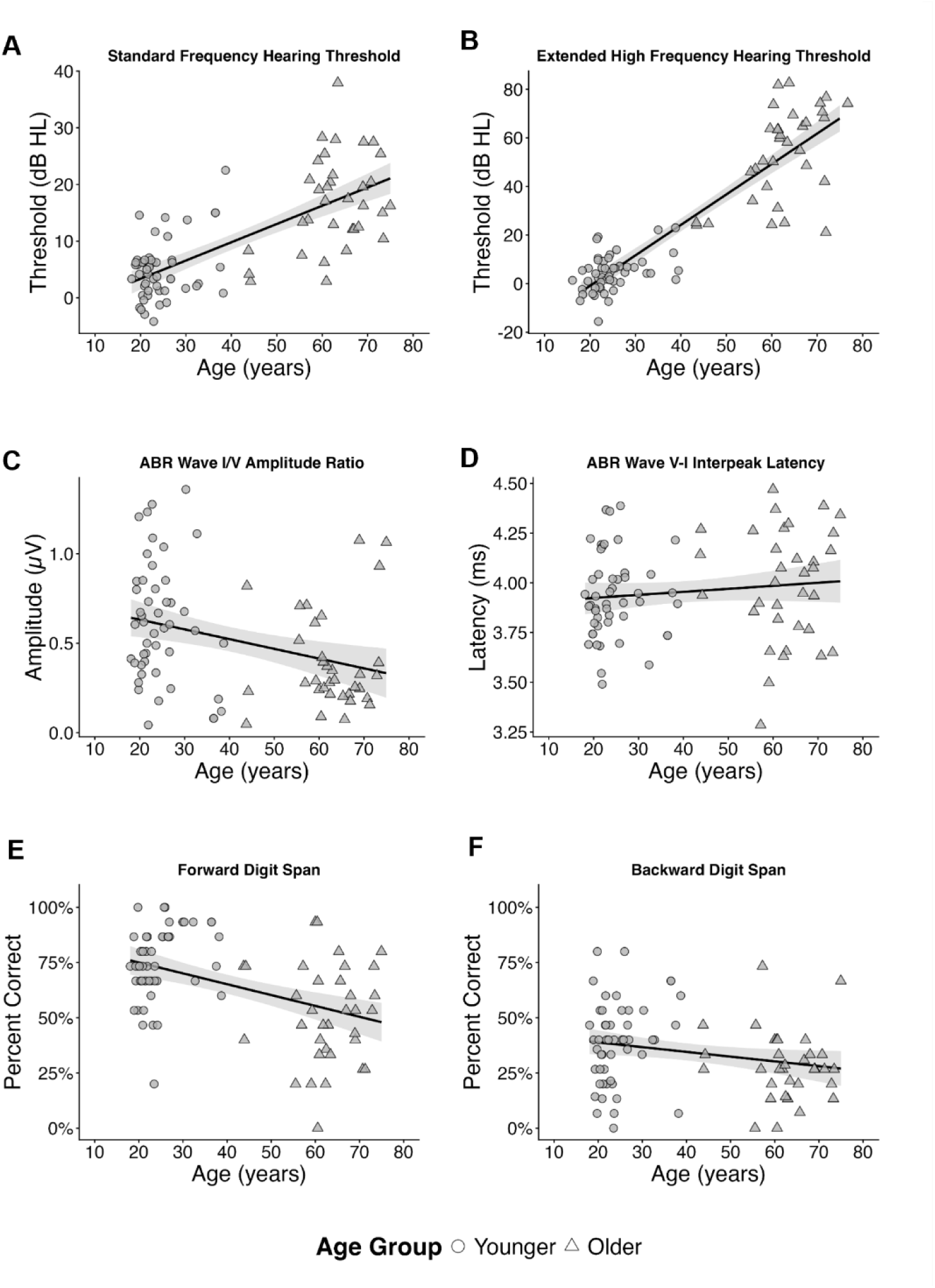
Relationships between predictor variables (y-axes) and participant age in years (x-axes). Predictor variables include A) hearing thresholds at standard frequencies, B) hearing thresholds at extended high frequencies, C) auditory brainstem response (ABR) wave I/V amplitude ratio, D) ABR wave V-I interpeak latency, E) forward digit span task scores, and F) backward digit span task scores. The line on each plot indicates the line of best fit, with the shaded area around the line representing the 95% confidence interval. Circles represent data from individual younger adults and triangles represent data from individual older adults.

## 4. Discussion

Acoustic cues help listeners segregate target speech from background noise, a crucial ability for accurate speech-in-noise recognition. In this study, younger and older adults completed a speech-in-speech perception task that emphasized the use of acoustic cues to investigate how well individuals of different ages can rely on F0 and ITDs to attend to the target talker. Individual participant scores differed between the Pitch and Spatial cue conditions (see Figure 4) and stepwise regression analyses indicated distinct patterns of predictor variables for these speech-in-speech perception task conditions. These findings provide evidence that individual differences in use of F0 and ITDs for speech stream segregation contributed to participants’ scores on the task. In addition, scores on all three conditions were significantly lower for the group of older participants compared to younger participants. This result demonstrates that age-related factors influence acoustic cue use for speech recognition in background noise.

### 4.1 Speech-in-Speech Recognition Performance

Overall, and when data were separated by age group, participants achieved the highest scores in the Pitch condition, suggesting that F0 was the most salient cue for speech-in-speech perception. Many previous studies have demonstrated benefits in speech recognition performance when F0 differed between target and competing speech. For example, Assmann (1999) found that keyword recognition scores for high-predictability sentences improved by 23% when the difference between target and competing speech increased from zero to eight semitones. Prior investigations have shown substantial score improvements on speech-in-speech recognition with as little as a two- or three-semitone difference between target and competing speech (e.g., Bird & Darwin, 1998; Brokx & Notebloom. 1982; Darwin et al., 2003). In the current study, the target was separated from distractors by 6 semitones, a relatively large difference between talkers (as is common in everyday listening scenarios). The six-semitone difference between target and masker used in the current study was therefore large enough to aid participants in target speech identification.

Participants received the lowest scores on average for the Spatial condition, indicating that the ITD cue was most difficult to use to segregate the target from the maskers. Spatial cues were limited to ITDs in this study to determine the effects of age and sensory factors on participants’ ability to use this specific cue for speech-in-speech perception. However, listeners use interaural intensity differences (IIDs) and spectral cues in addition to ITDs for sound localization in everyday listening scenarios. ITDs alone are thus an “unnatural” cue to rely on solely for spatial localization of a target speech stream. For example, Deng et al. (2019) tested young adults on a similar speech-in-speech recognition task, in which participants identified /ba/, /da/, and /ga/ syllables from a target stream in the presence of two distractor streams. Spatial separation between target and distractors was created by using either individualized head related transfer functions (HRTF), individualized IIDs, or a +/- 300 µs ITD (as in the current study).

Deng et al. (2019) found that listeners recalled phonemes from the target speech stream less accurately in the ITD condition relative to the conditions with more natural, individualized spatial cues. In addition, the score range for the ITD condition found in that study was similar to that observed for young adults in the current study, suggesting the Spatial cue condition performance may have indeed been higher if HRTFs or other individualized spatial cues were used instead of ITDs.

In addition, while the “eeeee” cue for pitch of the target talker was played at the beginning of each trial for all conditions, presentation of this sound was not useful for the Spatial condition. In the Pitch and Both Cue conditions, in which attention to this cue was indeed beneficial to participants, the “eeeee” presentation may have provided an initial sound on which listeners could build stream perception. For example, Bregman & Rudnicky (1975) showed that presenting precursor tones that were a similar frequency to tones in a target stream facilitated perception of the target stream. The lack of a perceptual scaffolding opportunity in the Spatial condition could provide an additional potential explanation for the significantly lower scores in that condition compared to the Pitch and Both Cue conditions.

Surprisingly, average performance on the Both Cue condition was lower than performance for the Pitch condition. Although we expected the availability of both F0 and ITDs to be beneficial to participants’ performance, it may have instead been detrimental. The Both Cue condition may have unintentionally elicited attention switching, in which participants constantly switched their focus between cues, thereby increasing the processing resources required for the task. Evidence to support this theory was revealed by analyses examining effect of cue condition on task scores for each age group. Only the older adult group demonstrated significantly lower scores on the condition with both cues available relative to the Pitch condition (see Figure 3). Performance on the Pitch and Both Cue conditions were statistically similar in young adults, indicating that while the addition of the ITD cue was not necessarily helpful, it also did not detract from task performance in the younger group. Reduced cognitive-perceptual processing occurs with age (see Pichora-Fuller, 2003 for review) and hearing loss can decrease cued auditory attention in competing talker scenarios (Caso et al., 2024; Dai et al., 2018). It is therefore likely that the Both Cue condition involved additional processing demands relative to conditions containing just one cue, potentially explaining why the inclusion of both cues for speech-in-speech recognition increased task difficulty for older adults.

### 4.2 Age Effects on Acoustic Cue Use for Speech-in-Speech Identification

Compared to younger adults, older adults on average achieved significantly lower scores on all three cue conditions of the speech-in-speech recognition task. Previous research has demonstrated poorer F0 (Moore & Peters, 1992; Vongpaisal & Pichora-Fuller, 2007; Souza et al., 2011) and ITD (e.g., Strouse et al., 1998; Grose & Mamo, 2010) discrimination in older adults compared to young adults. Results from the current study extended these findings from basic sensory discrimination tasks to functional use of F0 and ITDs for speech-in-speech recognition. Our results are also in line with prior studies that have demonstrated reduced spatial release from masking in older relative to younger adults. Gallun et al. (2013) and Zobel et al. (2019) both found that young and older adults benefited from spatial separation between target and masker speech, but older adults experienced significantly less spatial release from masking, even when controlling for hearing thresholds at standard frequencies. Dubno et al. (2002) similarly found that older adults with normal hearing at standard thresholds demonstrated greater spatial release from masking than older adults with hearing loss, but both older participant groups achieved lower spatial release than the young adult group. Those investigations all used speakers to create spatial separation between target and masker speech; the similar findings from the current study, which simulated spatial separation via ITDs presented through earphones, further emphasize the importance of ITDs for horizontal plane sound localization (e.g., Wightman & Kistler, 1992).

The observed differences between younger and older adults on the speech-in-speech perception task may have been driven by sensory coding of the acoustic cues available in each condition. Hearing thresholds (at standard frequencies or EHFs) significantly predicted performance in all cue conditions; the older adults in our study did demonstrate higher average thresholds than younger adults (see Figure 7A and 7B) and reduced cochlear function likely contributed to group differences in performance. In addition, and not assessed here, older adults demonstrate a decline in neural representation of both fine timing and of frequency that affects coding of acoustic cues. For example, Clinard et al. (2010) and Heidari et al. (2018) found weaker electrophysiological frequency following responses (FFRs) with age in adults with normal hearing thresholds through 4000 Hz. Ross et al. (2007) and Ozmeral et al. (2016) both found reduced cortical representation of ITDs in older adults relative to young adults. Anderson et al. (2011) additionally found that older adults with poorer encoding of F0 at the brainstem level also performed more poorly on a test of speech-in-noise recognition, demonstrating a link between physiological coding of an acoustic cue and one’s ability to hear in noise. Results of the current study add to prior evidence that age-related physiological deficits cause difficulties in using acoustic cues to perceptually separate competing talkers.

While older adults as a group demonstrated significantly poorer performance on the speech-in-speech recognition task relative to younger adults, participants in each age group demonstrated a wide range of scores on each cue condition (see Figure 3). Participants also varied in the difference in scores between the Pitch and Spatial conditions (see Figure 4A and B). Although this study primarily investigated the relationship between sensory coding and functional use of acoustic cues, individual differences in perceptual compensation may have also played a role (as in Winn et al, 2013; Winn & Livosky, 2015). For example, the poorest performing older adults for the Pitch condition achieved higher scores on the Spatial condition (see Figure 4D), suggesting that they may compensate for diminished ability to use pitch cues by relying more on spatial cues in everyday listening. Future work could examine cue trading between pitch and spatial cues to further investigate the relationship between older adults’ sensory processing and auditory perceptual strategies.

It is important to note that the “older” group in this study included participants aged 40 and older, suggesting that reduced ability to use acoustic cues for speech-in-noise understanding begins in middle age. Prior studies that have found age-related declines in auditory system function in individuals as young as 40 (Jerger and Hall, 1980; Kumar, 2022; Zink et al., 2025) which may contribute to decreased ability to perceive and utilize important cues for speech stream segregation. However, most participants in the older age group in the current study were 55 and older, and additional work is therefore necessary to investigate acoustic cue utilization in middle-aged individuals. Nevertheless, the current study builds on recent work of speech processing in middle-aged adults (e.g., Zink et al. 2025; Guo et al., 2025) and provides further evidence that age-related investigations of auditory perception should include both middle-aged and senior participants.

### 4.3 Factors that Best Predicted Scores on Each Cue Condition

The variables that best predicted performance on the speech-in-speech perception task differed between the cue conditions. For the Pitch condition, but not the Spatial or Both conditions, hearing thresholds at standard frequencies explained a significant proportion of variance in speech-in-speech identification scores. This finding is consistent with previous studies that have found listeners with hearing loss to achieve less benefit from F0 differences between target and masker relative to normal hearing listeners. Kidd et al. (2019) found that, relative to normal hearing individuals, young adults with sensorineural hearing loss demonstrated less release from masking when target speech and maskers differed in F0. Thresholds on the task also became poorer with increasing hearing loss in their study. Mackersie et al. (2011) applied individualized spectral shaping and amplification to task stimuli but still found that older listeners with hearing loss benefited less from target and masker F0 differences. The relationship between hearing thresholds and use of F0 for stream segregation is therefore not purely an issue of audibility but is also due to the reduced frequency selectivity and diminished temporal fine structure processing that occur with outer hair cell loss – as both decrease pitch coding.

Interestingly, although hearing thresholds at standard frequencies significantly predicted Pitch cue condition scores in the current study, all younger participants had normal hearing thresholds and older participants had only mild to moderate loss, even at 8000 Hz. These findings demonstrate that cochlear function is a primary contributor to F0 cue perception even in listeners without significant hearing loss.

Audiometric thresholds at EHFs (10k and 12.5k Hz) were present in the best fitting model for scores in the Spatial and the Both Cue conditions. These conditions both contained the ITD cue. In everyday listening, ITDs are most useful at low frequencies; the finding that EHF thresholds significantly predicted use of the ITD cue for speech-in-speech recognition is therefore surprising and may seem counterintuitive. One possible explanation for this result may stem from the relationship between EHF hearing thresholds and participant age. The predictor variable of EHF hearing was most strongly correlated with age (see Table 2 and Figure 7).

Rather than being a direct predictor of ITD cue use, a common aging mechanism may impair both EHF hearing and ITD perception. Another potential explanation is that elevated EHFs may reflect upstream neural pathology that could disrupt ITD coding, as has been found in some previous research (e.g., Shaheen et al., 2015; Mehraei et al., 2017; Parthasarathy & Kujawa, 2018; Budak, 2022), but not observed in other studies (e.g., Grose et al., 2017; Prendergast et al., 2017; Prendergast et al., 2019; Patro et al., 2021; Patro et al., 2025). Additionally, individuals with hearing loss at standard frequencies demonstrate poorer ITD perception (Dai et al., 2018); participants in the current study only had mild to moderate loss at standard frequencies, and therefore EHF thresholds may have better reflected damaged cochlear processes that decrease ITD perception in this population. The relationship between EHF hearing and ITD processing therefore warrants future investigation.

Still, previous research has found that frequencies above 8 kHz, in addition to sufficient audibility of that high frequency information, are important for localizing sound (Best et al., 2005; Brungart & Simpson, 2009). ITDs were the only spatial cue available for localizing target speech in the spatial condition, and it is possible that poorer high-frequency hearing decreased general spatial localization ability. Further, prior studies have found that individuals with elevated EHF thresholds self-report greater difficulty hearing in background noise (Motlagh Zadeh et al., 2019; Saxena et al., 2022) and achieve lower scores on spatialized speech-in-noise recognition tasks than those with better EHF hearing (Çolak et al., 2024). Although these findings might be due to reduced audibility of spectral cues at very high frequencies, results from the present study suggest that decreased ability to use ITD cues for spatial localization of a target talker could be one link between elevated EHFs and poor ability to hear in noise.

Normalized metrics of ABR wave amplitude and latency did not significantly predict performance on any cue condition of the speech-in-speech identification task in the expected direction. Some previous studies have found correlations between one or more ABR metrics and scores on a speech-in-noise perception task (Bramhall et al., 2015; Valderrama et al., 2018), but many have not observed this relationship (Guest et al., 2018; Prendergast et al., 2019; Mussoi, 2026). A possible explanation for these null results is that ABRs are not appropriate for predicting perception of complex stimuli. ABRs are a measure of neural response strength and signal transmission timing but may not be indicative of the neural precision that is required for accurate acoustic cue encoding. A measure such as the frequency following response (FFR), envelope following response (EFR), or frequency modulation following response (FMFR) better represent the relationship between auditory subcortical function and use of acoustic cues, as these metrics assess auditory phase-locking to acoustic features rather than synchronous neural onset activity. Studies using these neural assessments have shown that individuals with more precise subcortical coding perform better on speech-in-speech perception tasks in which target talkers and competing talkers differed in spatial location (via ITDs; Bharadwaj et al., 2015; Ruggles et al., 2011) and in F0 (Parthasarathy et al., 2020). Individual differences in neural synchrony may better predict one’s ability to use acoustic cues for segregating speech from competing speech than do the neuronal population responses measured by ABR.

However, it should be noted that all electrophysiological metrics of auditory processing are confounded by hearing thresholds (e.g., Bharadwaj et al., 2019). Of particular relevance to the current study, EHF hearing thresholds (representing basal cochlear function) have been found to influence ABR wave I amplitude even in adults with normal hearing thresholds through 8 kHz (Patro et al., 2024; Sendesen et al., 2026). It is difficult to assess neural function independent of cochlear function in humans, further complicating examination of the relationships between auditory neural processing and use of acoustic cues for speech-in-speech perception.

Backward digit span task scores, a measure of working memory capacity and executive control, significantly predicted scores on the Spatial and Both Cue conditions. As these two conditions were the most difficult, particularly for older adults (see Figure 3), this finding suggests that at least that group of participants relied more heavily on cognitive ability when the task was more challenging. Forward digit span task scores were not a significant predictor of any condition, indicating that greater cognitive resource allocation, not just greater memory span, facilitated performance on the speech-in-speech recognition task. This finding is in line with other previous studies that have found a relationship between speech-in-noise perception and scores on other tests of working memory capacity and executive function (Stenbäck et al., 2021; Gordon-Salant & Cole, 2016; Yeend et al., 2017) attention (Yeend et al., 2017), and inhibitory control (DiNino et al., 2025). Backward digit span task scores were also present in the best-fitting model for the Pitch condition but only trended toward being a significant predictor (see Table 2). Still, the finding of a positive relationship between backward digit span scores and performance on all three conditions of the speech-in-speech recognition task suggests that poor cognitive function may limit performance on auditory tasks regardless of the acoustic content of the stimuli.

### 4.4 Study Limitations

As with any investigation, this study had several limitations that should be addressed. First, discrimination data of F0 and ITDs were not collected to compare to participants’ scores on each cue condition of the speech-in-speech recognition task. A direct link between poor cue discrimination and reduced ability to utilize those acoustic cues for speech stream segregation could therefore not be established. Second, ABRs were used to assess subcortical auditory processing due to their common use in clinical settings, but metrics of auditory neuron phase-locking such as EFRs or FFRs may have been better predicted perceptual use of acoustic cues.

Lastly, the stimuli used in the speech-in-speech recognition task (/ba/, /da/, and /ga/) were chosen because they limit linguistic contributions to task performance, placing greater emphasis on basic sensory processing to perform the task. However, stimuli such as sentences or even audiobook stories would be more representative of everyday listening scenarios. Future research could expand this work using more ecologically valid stimuli.

## 5. Conclusion

Previous research has commonly linked age-related changes in auditory sensory processing to older adults’ challenges hearing in noise (e.g., Anderson et al., 2011; see Pichora-Fuller & MacDonald, 2009 for review). Evidence from the current study suggests that a decline in auditory function with age decreases one’s ability to use the important acoustic cues for segregating target sounds from background noise, thereby impairing speech-in-noise perception. This study provides one potential connection between altered auditory processing with age and older adults’ difficulties hearing in noise (e.g., Plomp & Mimpen, 1979; Humes, 1996 for review). The current study also builds on findings from prior studies that found poorer F0 and ITD discrimination in older relative to younger adults. Evidence from this study suggests that decreased perception of those cues with age extends to functional use of those cues for segregating speech, demonstrating that reduced sensitivity to these acoustic features has consequences for speech understanding in challenging listening environments.

The speech-in-speech perception task in the current study emphasized auditory selective attention - forming auditory objects to attend to one talker and ignore others - which helps listeners follow conversations in real-world “cocktail party” scenarios (Shinn-Cunningham, 2008). The task used in this study required listeners to use acoustic cues to segregate target speech from competing maskers, a process that is critical for accurate speech-in-noise perception in everyday listening. Still, future work might examine whether the same sensory and cognitive factors that predicted performance in the speech-in-speech perception task used in this study extend to more complex, naturalistic listening situations, such as multi-talker environments with reverberation or the perception of linguistically complex speech, which more accurately represent everyday listening environments and in which the demands on acoustic cue processing may be even greater. Additionally, given the evidence for neural changes beginning as early as middle age, future investigations would benefit from including individuals aged 40+ adults to better characterize the trajectory of these perceptual declines before they become clinically significant and/or to identify potential targets for early audiological intervention.

## Acknowledgements

We would like to thank Kennidy Dyer-DeCator, Mythili Thamilchelvam, and Alanah Myszka for assistance with data collection. We would also like to thank the participants for their time and effort. This study was funded by an Emerging Research Grant from the Hearing Health Foundation.

## Data Availability Statement

The data from this investigation are hosted on Dryad and available here: https://doi.org/10.5061/dryad.4xgxd25s6

## Notes

### Competing Interest Statement

The authors have declared no competing interest.

### Author Declarations

All study procedures were approved by the University at Buffalo Institutional Review Board.

### Summary of Updates

Text has been added/changed to clarify the goals of analyses and to provide better interpretation of results.

